# Network-based assessment of HDAC6 activity is highly predictive of pre-clinical and clinical responses to the HDAC6 inhibitor ricolinostat

**DOI:** 10.1101/2020.04.23.20066928

**Authors:** Tizita Z. Zeleke, Qingfei Pan, Cody Chiuzan, Maika Onishi, Mariano J. Alvarez, Erin Honan, Min Yang, Pei Ling Chia, Partha Mukhopadhyay, Sean Kelly, Ruby Wu, Kathleen Fenn, Meghna S. Trivedi, Melissa Accordino, Katherine D Crew, Dawn L Hershman, Matthew Maurer, Simon Jones, Andrea Califano, Kevin Kalinsky, Jiyang Yu, Jose Silva

## Abstract

Despite the anticancer activity of pan-histone deacetylase (HDAC) inhibitors, their clinical use has been limited due to toxicity. However, the development of more specific inhibitors that selectively inhibit individual HDACs is emerging as a novel and well-tolerated alternative. Here, we present the results of the first clinical trial evaluating the activity of ricolinostat (the leading HDAC6 inhibitor) in breast cancer (BC) patients.

We have developed a computational network-based algorithm to evaluate the activity of the HDAC6 protein, based on the enrichment of its transcriptional targets in differentially expressed genes (HDAC6 score). Through preclinical *in vitro* and *in vivo* studies, we confirmed that the HDAC6 score can stratify the sensitivity of BC cells to ricolinostat treatment and may thus have value as a predictive biomarker. Moreover, analysis of ∼3,000 primary human breast cancers showed that ∼30% of them present high HDAC6 scores. Based on these results, we designed a phase Ib clinical trial to evaluate the activity of ricolinostat plus nab-paclitaxel in metastatic BC patients. Study results showed that the two agents can be safely combined, that clinical activity is identified specifically in patients with HR+/HER2-disease, and that the HDAC6 score was predictive of response. Expansion of our analysis to other tumor types identified multiple cohorts enriched in high HDAC6 score samples. These results suggest that the HDAC6 score may provide an effective, CLIA certified predictive biomarker of ricolinostat sensitivity in multiple human cancers.

**SIGNIFICANCE:** The clinical use of HDAC inhibitors is hampered by the toxicity associated with blocking multiple HDACs. Here, we show that the specific HDAC6 inhibitor ricolinostat is safe and presents clinical activity in breast cancers and that the HDAC6 score has predictive biomarker potential to identify patients who can benefit from this therapy.

## INTRODUCTION

Homeostasis of cancer cells presents different oncogene and non-oncogene dependencies compared to non-transformed cells. Therapies aimed at targeting these dependencies represent more selective and less toxic anticancer strategies than standard chemotherapy (1). In this regard, inhibition of histone deacetylases (HDACs) using pan-inhibitors has proven anticancer activity, especially in hematopoietic malignancies (2). However, toxicity associated with pleiotropic inhibition of multiple HDAC family members has limited their clinical use (2). Thus, the interest has turned towards more selective inhibitors, targeting specific HDACs.

Recently, we reported that the viability of inflammatory breast cancers (IBC) depends on the function of histone deacetylase 6 (HDAC6) (3). HDAC6 is a class-IIB deacetylase responsible for deacetylating a variety of substrates. Although the full substrate repertoire is far from being fully characterized, HDAC6 has emerged as an essential component of the aggresome (4-6). We thus reasoned that other breast cancers (BCs) in addition to IBCs may present the same dependency and that identifying patient populations that can benefit from HDAC6 targeted therapy would be necessary for a rapid transition into the clinic. Thus, we first performed system biology studies suggesting a marked HDAC6 activity increase in HDAC6-dependent cells, where it was identified as a master regulator of cancer cell state, based on the enrichment of its regulatory targets in differentially expressed genes (3). Then, we developed an algorithm based on mRNA expression profiling (HDAC6 score) to quantify the activity of HDAC6 in individual tumor samples (3). Next, we used a variety of experimental models that includes *in vitro* cultures as well as *in vivo* mouse models to confirm the correlation between the HDAC6 score and the anticancer response to HDAC6 inhibition in BC cells. From these analyses, the HDAC6 score emerged as a candidate biomarker for the effective identification of tumors where HDAC6 behaves as a master regulator of tumor cell state, thus presenting a non-oncogene dependency essential for cancer cell viability.

In this study, we used the HDAC6 score to analyze all the primary tumors included in the TCGA and METABRIC data sets (∼3,000 primary cancers). Interestingly, we found that a group of BCs (∼30% of all BCs) that were enriched in hormone receptor-positive pathological and molecular characteristics (HR+) present an HDAC6 score predictive of potential response to HDAC6 inhibitors. Based on these results, we designed a phase Ib clinical trial in partnership with Acetylon/Celgene to investigate ricolinostat, the leading HDAC6 inhibitor (HDAC6i) (7) plus nab-paclitaxel as BC therapy (clinical trial ID#: NCT02632071). The latter was included based on pre-clinical data showing synergistic activity with ricolinostat, as well as for consistency with standard of care treatment in BC. Notably, we observed that ricolinostat plus nab-paclitaxel can be safely combined and that clinical activity is identified specifically in patients with HR+/HER2-disease. Furthermore, retrospective analysis confirmed that HDAC6 score analysis could successfully stratify patients based on clinical outcome, including based on the NY CLIA-certified OncoTarget test.

Finally, we also expanded these studies to all cancer cohorts in TCGA, using tumor type-specific HDAC6 scores, and confirmed their predictive values by dose-response studies *in vitro*. Overall, our studies support coupling of ricolinostat in combination with cytotoxic treatment with HDAC6 score for treating multiple human cancers.

## RESULTS

### Next-generation HDAC6 score predicts breast cancer sensitivity to HDAC6i

Recently, we reported that HDAC6 function is essential for maintaining the viability of a BC subtype called inflammatory breast cancer (IBC, ∼1-4% of all BC (8)) and demonstrated that the leading HDAC6 inhibitor ricolinostat (7) induces IBC cell death *in vitro* and *in vivo* (3). HDAC6 is rarely amplified or mutated and expression level of HDAC6 is similar between IBCs and non-IBC. However, system biology analysis based on transcriptomic profiling aimed at measuring protein activity (3) showed marked differential HDAC6 activity. In brief, we first used the ARACNe algorithm (9) to identify candidate HDAC6 transcriptional targets (i.e., transcripts whose expression is affected by HDAC6, HDAC6 regulon) from microarray-based transcriptome profiles of the TCGA breast cancer cohort. Then, we integrated the expression of all transcripts of the HDAC6 regulon in a single score, termed as HDAC6 score, by summarizing their expression values. Thus, the HDAC6 score represented the inferred HDAC6 activity (3).

Since our data revealed that dependency on the HDAC6 function was linked to high HDAC6 score in IBCs, we decided to explore its utility as a biomarker of HDAC6 inhibitor sensitivity. To accomplish this goal, we first assessed the fraction of breast tumor samples that would be predicted as HDAC6i based on the HDAC6 score analysis across both the BRCA-TCGA (10) and METABRIC (11) data sets (>3,000 primary tumors) as well as the set of BC lines available at the cancer cell line encyclopedia (CCLE; 47 lines) (12) (Supplementary Figure S1A).

While the original HDAC6 regulon used to generate the HDAC6 score was based on microarray-based gene expression profiling of 359 BRCA-TCGA samples (3), the inclusion of additional datasets including a more heterogeneous set of tumors and different profiling technologies (i.e., different gene expression microarray platforms and RNA-seq) required the adaptation of the original HDAC6 score. Thus, we revised the original regulon used to compute the HDAC6 score—i.e. its repertoire of positively regulated and repressed transcriptional targets— to make it consistent with these different technologies and tumor models. For this, we used the SJARACNe (13), an algorithm designed to reverse-engineer gene regulatory networks from large number of transcriptomics profiles (n > 100). This generated a refined HDAC6 regulon specific to breast cancer based on the integrative analysis of all available gene expression profiles from the BRCA-TCGA (RNA-seq, n=1,221) and METABRIC (microarray, n=1,904) cohorts. We also improved the HDAC6 score calculation by using the NetBID (data-driven network-based Bayesian inference of drivers) algorithm (14). As expected, despite marked profiling technology and sample preparation protocol differences between these datasets, HDAC6 activity predicted by the original and updated HDAC6 regulon were strongly correlated (*p* = 1.7E-33, Supplementary Figure S1B and Supplementary Table 1). Additionally, they captured known functions of HDAC6 such as unfolded protein response (Supplementary Figure S1C) and, consistent with previous findings, confirmed higher HDAC6 activity levels in IBCs (3) (Supplementary Figure S1D).

Using the updated regulon, we measured HDAC6 scores across all patient-derived samples and cell lines in the three data sets described above. Here, we observed that a significant fraction of all primary BCs ∼30% had higher HDAC6 scores than the median IBC HDAC6 score, suggesting potential sensitivity to HDAC6 inhibitors (Supplementary Figure S1D). Interestingly, high HDAC6 scores were not equally represented across BC subtypes but rather significantly enriched in the HR+ and HER2+ clinical subtypes (Figure 1A) and in the luminal-B and HER2-enriched molecular subtypes (Supplementary Figure S1E). This association was also evident in cell lines (Figure 1B).

**Figure 1.**
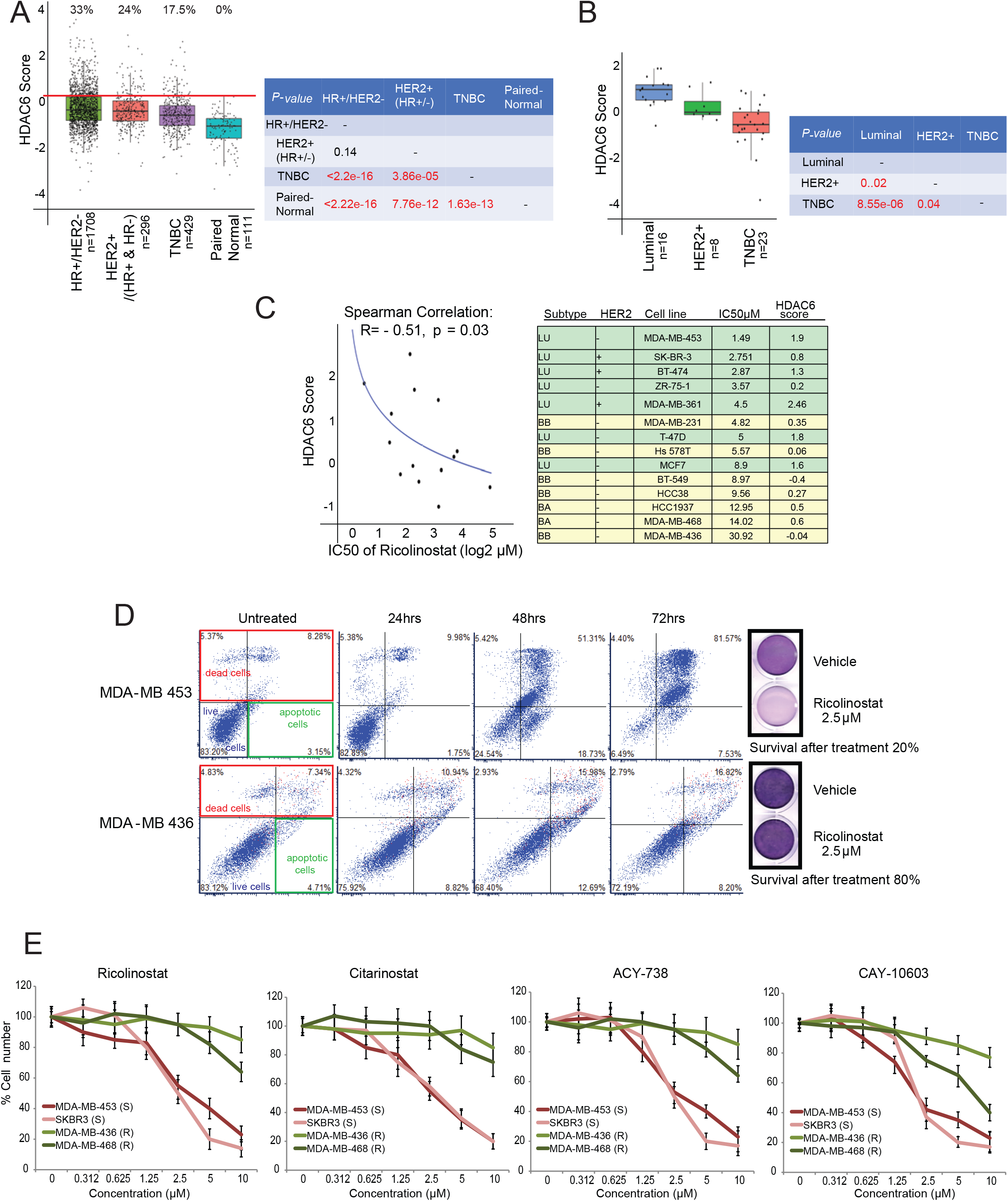
The HDAC6 score identify breast cancers sensitive to the HDAC6 inhibitor ricolinostat. (A) HDAC6 scores of all BCs from TCGA and METABRIC divided into clinical subtypes (HR. The red line represents the median of the HDAC6 scores in IBC samples and the numbers over each whisker plot indicate the percentage of samples over this value in each clinical subtype. The p-values comparing the HDAC6 scores of all the groups are also shown. (B) HDAC6 scores of all BCs cell lines in the CCLE data base divided into molecular subtypes. The p-values comparing the HDAC6 scores of all the groups are also shown. (C) Graphical representation and an associated table showing the correlation between HDAC6 score and response to ricolinostat. (D) Annexin-V/PE staining comparing the apoptotic response after Ricolinistat treatment of sensitive (MDA-MB-453) vs resistant (MDA-MB-436) BC cells. (E) Growth curves of sensitive (S) vs resistant (R) BC cells treated with four different HDAC6 inhibitors.

Next, we evaluated the correlation between the HDAC6 score and ricolinostat sensitivity in 14 BC lines representative of the major molecular subtypes (15), covering the full spectrum of HDAC6 scores (inferred from RNA-seq data from the CCLE). Specifically, we generated dose-response curves (8 doses ranging from 0 to 30μM) to measure cell line specific IC_50_ values. The analysis revealed strong association between HDAC6 score and sensitivity (*R* = −0.51, by Spearman’s correlation, *p* = 0.03), thus showing that cancer cell lines with high HDAC6 score present higher ricolinostat sensitivity on average (Figure 1C).

We had previously shown that ricolinostat treatment induces apoptosis on sensitive IBC cancer cell lines (3). To investigate if a similar mechanism is responsible for the anti-proliferative effect of the drug in non-IBC cells we first selected two lines with either low (MDA-MB-453) or high (MDA-MB-436) IC_50_ from the list. Next, we identified the lowest concentration of ricolinostat that induces robust accumulation of acetylated α-Tubulin—a well-known HDAC6substrate, without affecting accumulation of acetylated histones—established markers of pleiotropic class-I HDAC inhibition (i.e. off-target effects) (Supplementary Figure S2A). Annexin-V staining following ricolinostat treatment with that concentration of ricolinostat (2.5μM) revealed progressive accumulation of apoptotic MDA-MB-453 cells (sensitive), with only very minor effects in MDA-MB-436 cells (resistant) (Figure 1D).

To further demonstrate the specificity of the effect we generated IC_50_ values for 3 additional HDAC6 specific inhibitors in BC cells presenting the highest (MDA-MB-453 and SK-BR-3) and lowest (MDA-MB-436 and MDA-MB-468) sensitivity to ricolinostat. These inhibitors include citarinostat—a more soluble analog of ricolinostat (3, 16)—and two next-generation inhibitors that are structurally unrelated with ricolinostat, ACY-738 (17) and CAY10603 (18) (Supplementary Figure S2B). These analyses confirmed that all HDAC6 inhibitors reduced growth of ricolinostat-sensitive cell lines with only minor effects in ricolinostat-resistant ones, at the same doses (Figure 1 E).

### Ricolinostat shows anticancer activity *in vivo* on cancer cells selected by HDAC6 score analysis

Next, we investigated the anticancer activity of ricolinostat in BC cells *in vivo*. While these data show ricolinostat’s anticancer activity as a single agent, achieving clinically-relevant response *in vivo* generally requires combining targeted therapy with standard chemotherapy. Primary chemotherapy for BCs typically includes the use of anthracyclines and taxanes (19, 20). Thus, we decided to assess the therapeutic value of combining paclitaxel and doxorubicin with ricolinostat. For this, we used two commonly used methods: Isobole curve (21) and combination index (by Chou-Talalay equation) (22). These two analyses showed that both chemotoxic agents synergize with ricolinostat, in ricolinostat-sensitive MDA-MB-453 cells (Figure 2A), but not in ricolinostat-resistant MDA-MB-436 cells (data not show).

**Figure 2.**
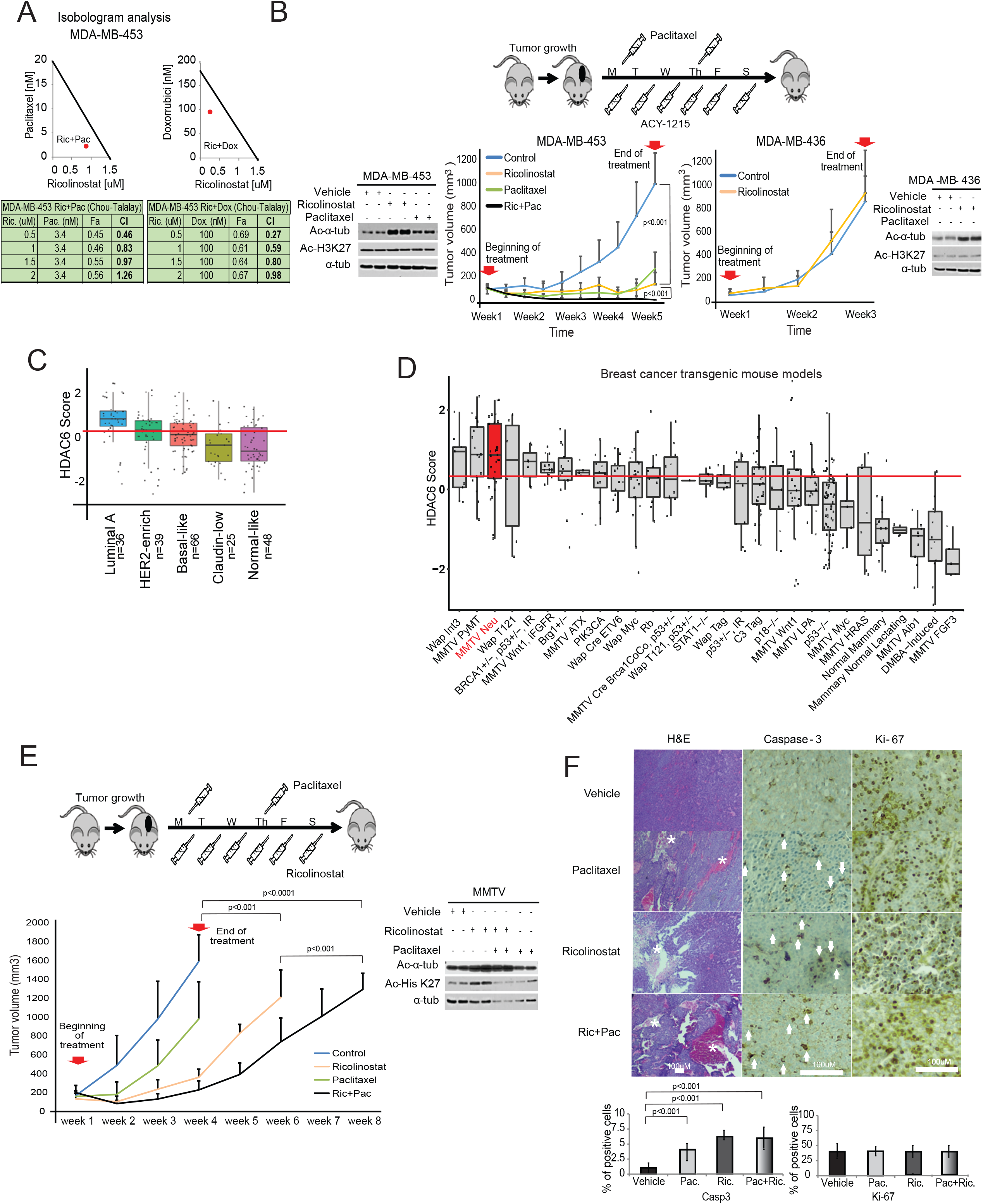
Anticancer activity of ricolinostat *in vivo*. (A) The graphic shows the synergistic activity between ricolinostat and commonly used chemotherapy (paclitaxel and doxorubicin). (B) Treatment of ricolinostat sensitive (MDA-MB-453) and resistant (MDA-MB-436) growing as xenografts in SCID mice. The cartoon illustrates the treatment regimen. The combinatorial effect with paclitaxel was also investigated in sensitive cells. On resistant cells, only ricolinostat was used because no effect was observed with the combo *in vitro*. The western blots show the accumulation of acetylated tubulin in tumors treated with ricolinostat. Additionally, the absence of off-target effect in class-I HDACs is shown by the minimum changes seen on the levels of acetylated Histone-3-K27 (two independent tumor samples are shown). (C) HDAC6 scores in tumors emerging in transgenic mouse models that recapitulate the molecular characteristics of human BCs. (D) HDAC6 scores in tumors emerging in 27 different transgenic mouse models of BC (with and without molecular characteristics of human BCs). In C and D the red line represents the mean of the HDAC6 scores in IBC samples. (E) Treatment of BC tumors emerging in the MMTV_Neu transgenic mouse model. The beginning and end of treatment are indicated by the red arrows. The cartoon illustrates the treatment regimen. The combinatorial effect of ricolinostat plus paclitaxel was also investigated. The western blots indicate the same that in panel B. (F) Histological intratumor evaluation of H&E, Caspase-3 and Ki-67 in tumor samples from panel E. Quantification is also shown in bar graphs.

To transition our studies with MDA-MB-453 and MDA-MB-436 to an *in vivo* context these cells were injected as mouse xenografts in the flanks of γ-SCID mice. After the tumors grew to ∼0.3cc, they were randomly assigned to one of several therapeutic regiments, including ricolinostat and paclitaxel as single agents and ricolinostat plus paclitaxel in combination (combo). Paclitaxel was eventually selected over doxorubicin because it is widely used for BC standard of care. Confirming *in vitro* results, ricolinostat demonstrated significant antitumor growth activity as a single agent in MDA-MB-453 but not in MDA-MB-436 xenografts. WT-blot measuring Ac-α-Tubulin and Ac-His-K27 in tumor extracts from treated animals confirmed that the effect was associated with specific HDAC6 inhibition, with minimal effect on other class-I HDACs. Interestingly, while a small tumor mass was still detectable in the sensitive cells at the end of the treatment period with ricolinostat (1 month), combination treatment with paclitaxel induced complete response (Figure 2B). Intratumoral evaluation of the treated animals showed that the ricolinostat response in MDA-MB-453 tumors was associated with higher apoptosis levels (activated Caspase-3) while no such effect was seen in ricolinostat resistant MDA-MB-436 cells (Supplementary Figure S2C).

To complement our preclinical *in vivo* studies with more pathophysiologically-relevant models, we evaluated response to ricolinostat in spontaneous tumors using transgenic mouse models. Numerous murine models of BC have been created to mimic the genetic aberrations found in human tumors. In particular, gene expression profiles of 385 tumors representative of 27 different genetically-engineered mouse models (GEMMs) of BC have been described and compared with human counterparts (23) (Supplementary Figure S2D). Consistent with the human tumor studies, HDAC6 score analysis using these murine gene expression profiles confirmed that mouse models recapitulating the molecular characteristics of human luminal breast cancers presented with the highest HDAC6 scores (Figure 2C). Since MMTV-Neu models tend to generate a homogeneous group of Luminal tumors (23) (Figure 2D and Supplementary Figure S2D), we investigated their response to the same ricolinostat regiments used to treat the MDA-MB-453 xenografts. Remarkably, when comparing single-agent treatments, we observed that ricolinostat alone was more effective than chemotherapy in MMTV-Neu tumors (Figure 2E and Supplementary Figure S2E). However, as also seen in MDA-MB-453 xenografts, combination of ricolinostat and paclitaxel produced the strongest antitumor effect. Consistently, intratumoral evaluation of treated animals showed that ricolinostat anticancer activity was associated with higher cell death levels with multiple areas of necrosis and apoptosis, and that this effect was stronger in the tumors treated with the drug combination (Figure 2F).

### Phase 1b clinical trial of ricolinostat combined with nab-paclitaxel in metastatic breast cancer

Taken together, these studies suggest significant association of ricolinostat sensitivity in BC with high HDAC6 score. To translate this hypothesis to a clinical context we designed an open-label phase Ib trial using ricolinostat in combination with nab-paclitaxel for patients with metastatic breast cancer (MBC). The primary objective of this study was to establish the safety, tolerability and identify the maximum tolerated dose (MTD) and recommended phase II dose (RP2D) of ricolinostat when combined with nab-paclitaxel. Additionally, the secondary objectives were to assess progression-free survival (PFS), overall response rate (ORR), clinical benefit rate (CBR) and to evaluate the HDAC6 score as a predictive biomarker by investigating the correlation between the HDAC6 score and clinical endpoints (Figure 3A).

**Figure 3.**
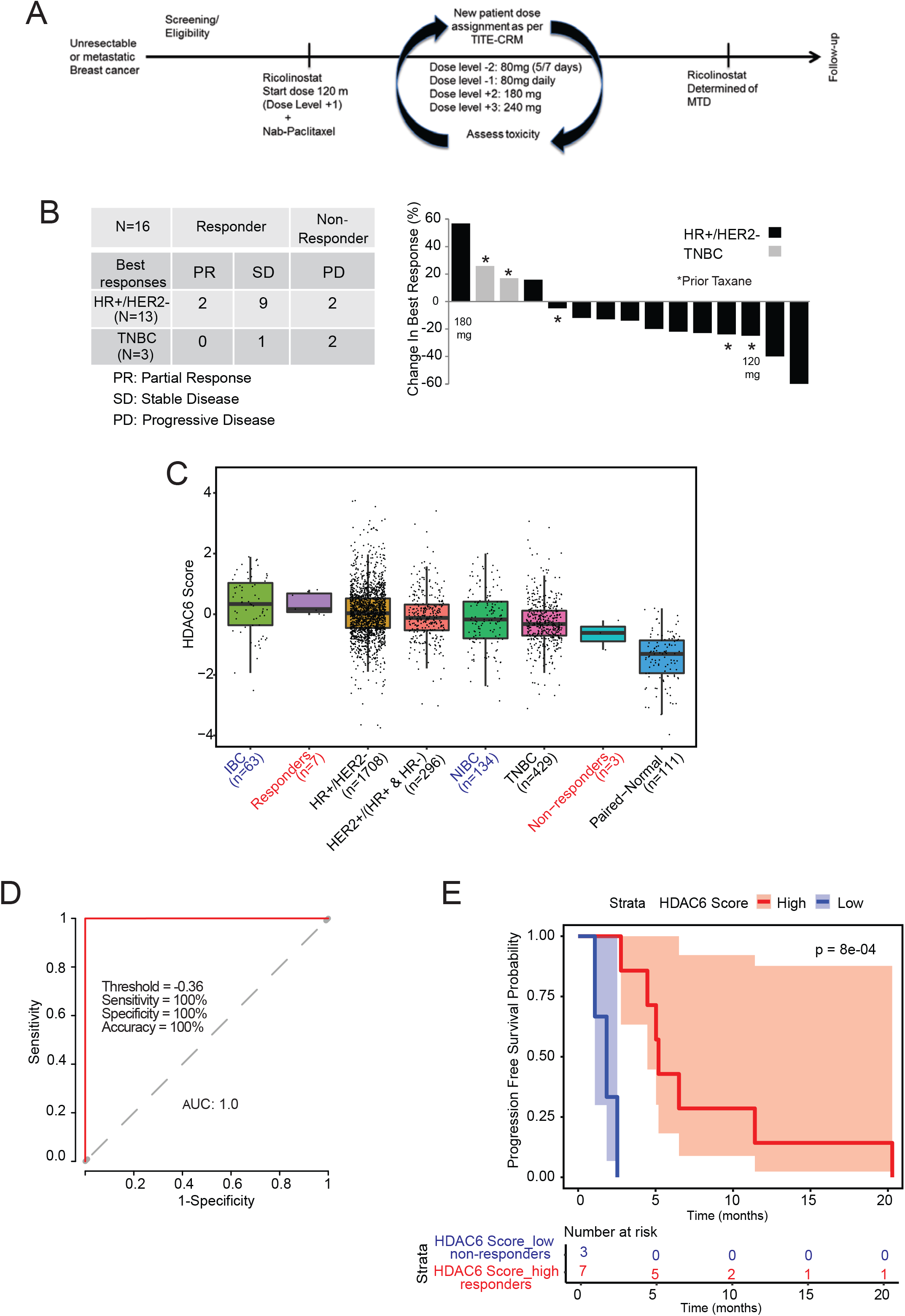
Phase 1b trial of ricolinostat combined with nab-paclitaxel in metastatic breast cancer. (A) Graphical description of the clinical study. (B) Waterfall plot showing the tumor best response for patients with measurable disease (right). The subtype characteristics of all patient with evaluable disease (n=16) is also shown (left). Note that one evaluable patient with stable disease did not have measurable disease and is not included in the waterfall plot (n=15). (C) The bar graph shows the HDAC6 scores in the patients in the trial (labeled in red) together with all the BC samples evaluated and separated by subtype. Labeled in blue are the IBC and the matched non-IBC series. (D) Receiver operating characteristic (ROC) curve plot for evaluation of HDAC6 score to predict the response of patients with breast cancer to ricilinostat from the clinical trial. The recommended cutoff of the HDAC6 score and corresponding sensitivity, specificity, and accuracy were inside the box. (E) Kaplan – Meier graphic showing the survival of the patients in the study separated by HDAC6 score (high/low= higher and lower than −0.36, the cutoff ofHDAC6 score based on the ROC analysis in the study).

In this trial, patients received ricolinostat orally (liquid) for 21 consecutive days of each 28-day cycle with nab-paclitaxel dosed at 100 mg/m2 on days 1, 8, and 15 until progression of the disease or unacceptable toxicity. Entry criteria included men or women with any metastatic BC subtypes. Measurable disease was not required. The MTD of ricolinostat with nab-paclitaxel was estimated via dose-escalation employing a time-to-event continual reassessment method (TITE-CRM) (24). The MTD was defined as the dose combination associated with a target probability of dose-limiting toxicity (DLT) of 0.25. The TITE-CRM used an empirical dose-toxicity model. A total of five pre-defined doses of ricolinostat were selected for the dose-escalation process Seventeen patients were accrued between March 2016 and February 2018. Of these, 16 patients had evaluable disease, as one patient dropped out at cycle 2 due to no longer wishing to participate in the trial and in the absence of any related toxicity. In the 16 evaluable patients, the median age was 57.5 years (range: 41-78), 14 were female (87.5%), 3 had triple-negative MBC, and 13 were HR+/HER2-MBC. The median number of prior lines was 3 (range: 0-10) (Supplementary Figure S3A). The first patient started at 120 mg/m^2^ qd, the second at 180 mg/m^2^ qd, and the remaining 14 patients were treated at 240 mg/m^2^ qd. No DLTs were seen in the DLT window of 8 weeks (first two cycles), and thus the MTD was not reached. Grade III events related to nab-paclitaxel included neutropenia (n=1), peripheral neuropathy (n=1), and 1 grade IV neutropenia. Grade III syncope related to ricolinostat was observed in 2 patients (Supplementary Tables 2-4). All of these events occurred after the DLT window. In the 16 evaluable patients, the following were best responses: 2 partial response (PR), 10 stable diseases (SD), and 4 progressive diseases (PD: 2 TNBC, 2 HR+/HER2-) (Figure 3B). All patients had measurable disease (Figure 3B), except for one evaluable patient without target lesions who was reported to have SD for 9 months. Three patients who previously received a taxane in the metastatic disease achieved SD with ricolinostat plus nab-paclitaxel. One patient with SD remains on treatment since Feb 2018 (17 months). The clinical benefit rate was 31.25%: 5/16 patients (2 PR + 3 SD > 6 months). All of these patients were diagnosed with HR+/HER2-MBC, except 1 stable disease with TNBC. Median PFS 5.3 months [95% confidence interval (CI): 4.45-11.0] (Supplementary Tables 5-7).

We were able to obtain tumor specimens in the form of paraffin sections (FFPE) with >50% in tumor content for 10 of the 16 evaluable patients (3 achieving PD and 7 showing SD or PR). RNA was obtained from these samples, subject to genome-wide RNA-seq and the expression profiles obtained were used to calculate the HDAC6 scores. Interestingly, when we compared the HDCA6-scores between patients showing PD (non-responders) and those with either SD or PR (responders), a statistically significant higher HDAC6 score was seen in responder patients (p=2.1E-3 Supplementary Figure S4A). HDAC6 score analysis integrating our clinical study with TCGA, METABRIC and IBC cohorts confirmed our previous hypothesis that patients with HR+/HER2-breast cancer respond better than those with TNBC (Figure 3C). Further, we used the receiver operating characteristic (ROC) curves (25) to characterize the sensitivity/specificity of the HDAC6 score using the trial data. Briefly, ROC curve analysis is a graphical plot that illustrates the diagnostic ability of a binary classifier system as its discrimination threshold is varied. The best cutoff value can be calculated by ROC analysis for continuous variables to predict dichotomous variables with the best sensitivity and specificity. In this analysis, the ROC curve analysis for the HDAC6 score achieved an area under the curve (AUC) of 1.0 (Figure 3D). Although the perfect AUC was likely influenced by the small number of patients, at a cutoff value of −0.36, the HDAC6 score gave rise to a clear separation between responders and non-responders with 100% accuracy, and it outperformed the subtype (HR+/HER2 or TNBC) that had an accuracy of 80% (2 out of 10 patients were mispredicted, including 1 HR+/HER2- and 1 TNBC). Finally, we examined the predictive power of HDAC6 score for patient prognosis, measured by PFS. We classified the patients into HDAC6 score high and low groups using the cutoff of −0.36 calculated by the ROC analysis. Patients with high HDAC6 score had a median PFS of 6.51 months (95% CI: 5.19-NA), which was significantly better (p = 8.0E-4, Figure 3E) than patients with low HDAC6 score who had a median PFS of 1.84 months (95% CI: 1.08-NA).

Transitioning any molecular biomarker to the clinic requires using a CLIA-certified test. The Darwin OncoTarget test, which is based ln the VIPER (Virtual Inference of Protein-activity by Enriched Regulon analysis) algorithm (26), was developed precisely to compute the activity of druggable proteins in cancer patients, based on the expression of their ARACNe-inferred transcriptional targets. The test has recently received CLIA certification from the NY State Dpt. of Health for the analysis of both fresh-frozen and FFPE samples and is available from the Department of Pathology and Cell Biology at Columbia (27). In particular, Darwing OncoTarget includes assessment of HDAC6 protein activity, among several other proteins for which a high-affinity inhibitor is available in the clinic. As a result, we assessed whether HDAC6 activity measured via this clinical-grade test was also predictive of patient sensitivity to the ricolinostat/nab-paclitaxel combination therapy in the Phase Ib study. Consistent with the results discussed in the previous sections, the HDAC6 score measured by Darwin OncoTarget was equally effective in stratifying patient sensitivity (*p* = 9.4E-3, Supplementary Figure S4B), achieving a classification of the 7 responders and 3 non-responders with a AUC = 0.9 [95% CI: 0.68-1.0] based on ROC analysis (Supplementary Figure S4C).

### Cancers with high HDAC6 score are found in a large variety of human cancers

Since our studies confirmed the prognostic value of the HDAC6 score in human patients, we decided to systematically assess the HDAC6 scores across a large repertoire of human primary malignancies and cancer cell lines. Specifically, we analyzed >10,000 gene expression profiles, representing 32 molecularly-distinct human malignancies represented in the TCGA database (https://www.cancer.gov/tcga). First, we generated tumor-specific HDAC6 regulons using the same approach successfully tested in breast cancer and used them to calculate the HDAC6 scores for all TCGA samples in a cancer type–specific manner. As expected, the number of genes that overlapped among the different tumor types was highly significant, although tumor type-specific differences were also noticeable (Supplementary Figure S5A, and Supplementary Tables 8-9). Next, we aimed to investigate whether a correlation exists between the HDAC-6 scores and the response to therapy in other tumor types.. For this, we performed dose-response studies to assess ricolinostat IC50 in 58 additional cancer lines, representing 11 different tumor types. Notably, confirming the breast cancer-specific findings, a significant anticorrelation was detected between HDAC6 score and IC_50_ (R = −0.44, *p* = 5.2E-5) (Figure 4A; Supplementary Figure S5B and S5C, and Supplementary Table 10). Finally, we assessed the HDAC6 scores in 1156 different cancer cell lines available in the CCLE cohort, representing 20 tumor types (12), as well as in 32 primary tumors (TCGA database) (Figure 4B and 4C and Supplementary Table 11). This analysis showed that the vast majority of human cancers present a wide distribution of HDCA6-scores. Because of the correlation between the HDAC6 score and the response to ricolinosat these results suggested that a significant subset of patients, across multiple tumor types, may benefit from treatment with this HDAC6i.

**Figure 4.**
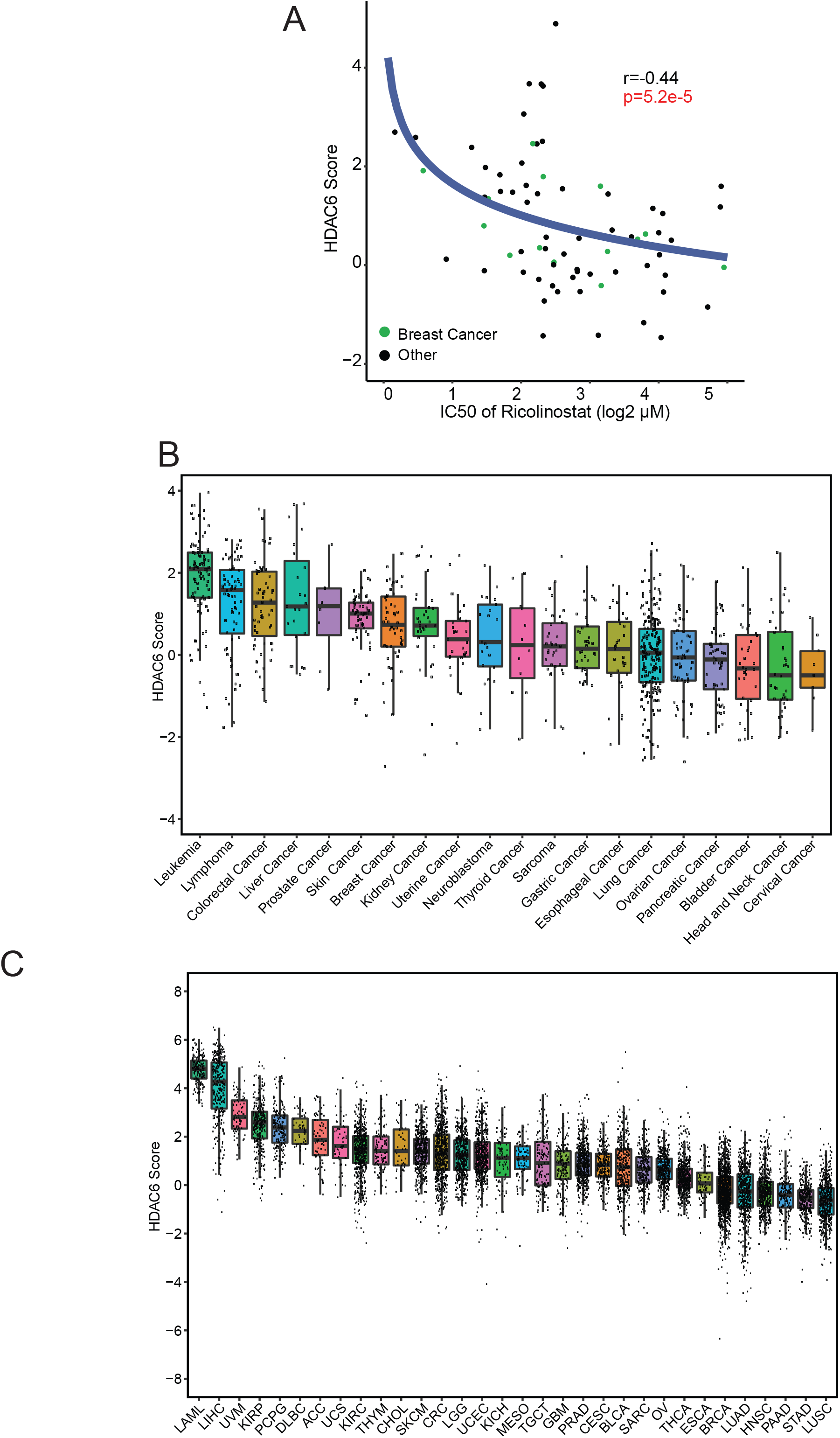
The HDAC6 score correlates with the response to ricolinostat in other cancer types. (A) Graphic showing the correlation between HDAC6 score and response to ricolinostat in 72 human cancer cell lines. (B) HDAC6 scores calculated for 1,156 different cancer cell lines available in the CCLE database representing 20 different human cancers and (C) for over 10,000 molecular profiles representing 32 different types of human cancer that have been collected in the TCGA database. LAML: Acute Myeloid Leukemia; LIHC: Liver Hepatocellular Carcinoma; UVM: Uveal Melanoma; KIRP: Kidney Renal Papillary Cell Carcinoma; PCPG: Pheochromocytoma and Paraganglioma; DLBC: Diffuse Large B-Cell Lymphoma; ACC: Adrenocortical Carcinoma; UCS: Uterine Carcinosarcoma; KIRC: Kidney Renal Clear Cell Carcinoma; THYM: Thymoma; CHOL: Cholangiocarcinoma; SKCM: Skin Cutaneous Melanoma; CRC: Colorectal Carcinoma; LGG: Brain Lower Grade Glioma; UCEC: Uterine Corpus Endometrial Carcinoma; KICH: Kidney Chromophobe; MESO: Mesothelioma; TGCT: Testicular Germ Cell Tumors; GBM: Glioblastoma; PRAD: Prostate Adenocarcinoma; CESC: Cervical Squamous Cell Carcinoma and Endocervical Adenocarcinoma; BLCA: Bladder Urothelial Carcinoma; SARC: Sarcoma; OV: Ovarian Serous Cystadenocarcinoma; THCA: Thyroid Carcinoma; ESCA: Esophageal Carcinoma; BRCA: BC; LUAD: Lung Adenocarcinoma; HNSC: Head and Neck Squamous Cell Carcinoma; PAAD: Pancreatic Adenocarcinoma; STAD: Stomach Adenocarcinoma; LUSC: Lung Squamous Cell Carcinoma.

## DISCUSSION

There are several key points related to the study presented in this manuscript that warrant further discussion. First, HDAC6 is a class-IIB member of the HDAC family, and thanks to the development of specific small molecule inhibitors, such as ricolinostat, is emerging as a promising therapeutic target. Anticancer activity of ricolinostat alone or in combination with additional drugs has been recently reported in preclinical models of multiple myeloma (MM) (7), pancreatic and ovarian cancer (16), esophageal cancer (28), melanoma (29) and lymphoma (30). Our studies presented here, showed that ricolinostat is also active in a variety of BCs enriched in HR+ and HER2+ characteristic.

Second, several early-stage clinical trials using ricolinostat in MM, lymphoid malignancies, leukemia, gynecological cancers and our own in BC are currently being evaluated. Because these are early-stage studies they have been mainly focused on describing the MTD of this HDAC6i alone or in combination with other drugs. Overall, and in agreement with our results, the clinical data from these studies confirmed that, at the range of doses used in our clinical trial, ricolinostat is safe, well-tolerated, and active (31-33).

Third, targeted therapies like ricolinostat represent exciting novel strategies to treat human cancers. Due to their ability to interfere with specific targets and pathways, they do not have the general undesired toxicity that is commonly seen with standard chemotherapy. However, these therapies do not have the wide spectrum of anti-cancer activity of the former therapies and generally need to be delivered based on the assessment of predictive molecular markers (34). As an example, anti-HER2 therapy such as trastuzumab or pertuzumab that are widely used to treat BCs are only active in the context of tumors that express high levels of this growth factor receptor. In fact, multiple specific inhibitors that have demonstrated efficient anticancer activity in some contexts have shown disappointing clinical results due to the lack of predictive biomarkers that can identify the correct patient population. Critically, the HDAC6 score developed in this manuscript provides an effective predictive biomarker to identify patients most likely to benefit from HDAC6i therapy. In terms of immediate clinical translation, we have shown that, consistent with prior clinical studies of drug sensitivity in multiple myeloma (35), the NY CLIA certified Darwin OncoTarget test produces HDAC6 scores that are highly predictive of patient sensitivity to ricolinostat plus nab-paclitaxel combination therapy.

Fourth, we have used the HDAC6 score to study a large variety of human cancer and found that cancer types such us AML (36, 37), lymphoma (30) and melanoma (29), where ricolinostat and other HDAC6 inhibitors have shown anticancer activity in preclinical models present the highest HDAC6 scores. Additionally, a fraction of the most common tumor types such as prostate, colorectal or B-cell lymphoma and some of the most deadly ones such as melanoma or glioblastoma were also at the top of the list. As our studies have also found a significant correlation between the HDAC6 score and the response to ricolinostat in a variety of cell lines other than BC, this opens the exciting possibility to use HDAC6i in multiple other tumor types. Combined with the availability of a clinical-grade test, these findings support the ability to perform additional HDAC6i clinical studies across multiple histologies and organ sites, thus improving the potential application of therapeutic drugs such as ricolinostat beyond BC in a variety of other tumors.

Our study has some limitations that are worth discussing. Although ricolinostat induces apoptosis in cancer cells *in vitro* and *in vivo*, the reasons for this are still unclear. Arguably, the main reason is the lack of characterization of the HDAC6 substrates and how acetylation influences their function. Only a few bonafide substrates such as α-Tubulin (6) and Hsp90 (38) have been well-validated and about over another half a dozen have been reported in the literature only a few times. Without this data, full understanding of the anticancer activity of HDAC6i is hampered. Nonetheless, cumulative evidence indicates that HDAC6 in mediating and coordinating various cellular events in response to different stressful stimuli (38-41). Thus, is likely than multiple pathways are involved in the anticancer activity of HDAC6i.

Finally, the ability to calculate the HDAC6 score for some of the patients recruited to our clinical trial study provides the first evidence that the HDAC6 score has predictive potential in the clinical setting. The ideal drug predictive biomarkers would integrate information of the intended target with potential mechanism of resistance and even off-target effects that can contribute to the final response. Because our study is a phase Ib the number of patients is still limited and the latest is not considered to calculate the HDAC6 score which may reduce its accuracy. Additional studies with larger cohorts are needed to further confirm these results and help to precisely define a threshold of HDAC6 score for patient stratification.

## Data Availability

The RNA-seq data from the clinical trial reported in this paper can be accessed at GEO: (GSE number GSE148623). The codes for the HDAC6 score calculation and other analyses are freely available at https://github.com/jyyulab/HDAC6-score.

## CONTRIBUTIONS

JS, JY and KK designed and coordinated the research and wrote the manuscript (JS coordinated the experimental preclinical studies. JY coordinated the computational analysis. KK coordinated the clinical trial). AC coordinated the Viper studies and wrote the manuscript.

TZ performed the preclinical experimental studies. QP performed the computational studies. Cody Chiuzan performed the statistical analysis of the clinical trial. MA performed the Viper studies. MY and SJ performed the dose response studies with ricolinostat. PC and PM collaborate with TZ performing animal studies. MO, MT, MA, SK, EH, RW, KF, KC, DH, MM and KK performed the clinical trial.

## ACKNOWLEDGEMENTS

We would like to thank members of the Silva and Yu laboratories for advice generating this manuscript and for technical assistance. Silva lab: Dayanira Alsina-Beauchamp and Rachel Werner; Yu lab: Xinran Dong, Koon-Kiu Yan, and Liang Ding. We also would like to thank Sui-Wan Lee from Mount Sinai’s CCMS core for technical assistance with mouse xenograft experiments. The authors also thank Keith A. Laycock, PhD, ELS, for scientific editing of the manuscript

This research was partially funded through the DOD Breakthrough award 151500 (JS), ALSAC (JY), the St. Jude Comprehensive Cancer Center Developmental Fund (JY), NIH R01 CA153233 (JS), NIH R01 CA153233 –supplement (TZ), R01 GM134382 (JY), U01 CA217858 (AC), S10 OD012351 (AC), and S10 OD021764 (AC), and the Irvin Scholar program (KK).

## MATERIAL AND METHODS

### Cell culture

All breast cancer cell lines were obtained from American Type Culture Collection (ATCC.

### Flow cytometry and cell viability

Cells were analyzed for phosphatidylserine exposure by annexin-V FITC / propidium iodide double staining using BD FITC Annexin V Apopotosis Detection Kit (Cat# 556547) according to the manufacturer’s instructions.

- ***Cell viability*** 5,000 cells/well were seeded in a 96 well white with clear bottom plates and treated with respective drugs for 72hrs and quantified for viability using CellTiter-Glo® Luminescent Cell Viability Assay (Cat# G7571). After treatment with drug was complete, media containing drug was removed and washed once with 1x PBS and replaced with equal volume of media and Cell Titer-Glo reagent and incubated for 5minutes on a platform rocker. Plates were then read using a SpectraMax M5 microplate reader at endpoint and luminescent setting.
- ***Drug synergy*** Using the same conditions and reagents as stated above, cells were treated with ricolinostat (0-20uM), paclitaxel (0-437.5nM) and doxorubicin (0-200nM) for 72 hrs to determine the dose curves. After validating 2.5uM of ricolinostat as the lowest concentration of the drug that induces robust accumulation of acetylated α-tubulin, the cells were treated with a combination of ricolinostat at 2.5uM and the same range of concentrations of paclitaxel and doxorubicin as indicated above for 72hrs and quantified for viability. The synergistic activity between ricolinostat and paclitaxel or doxorubicin was evaluated by two approaches: isobole curves and combination index using the Chou-Talalay equation.
- ***Cell viability by MTS*** Cells were plated in 96-well plates with 150 µl culture medium at an optimized cell density. Twenty-four hours later, testing compounds were added and the time zero plates were measured by MTS assay as G0 reference. Cell proliferation was measured by MTS assay after compound treatment for 3 days. Compounds dilution: 20mM stock solution of ACY-1215 in DMSO. On the day of treatment, compounds were freshly diluted from the stock solution to a working solution with 8 data points with final concentrations ranging from 0-30uM in culture medium.
- ***Crystal violet staining*** 50,000cells/well were seeded in a 24 well plate and treated with respective drugs for 72hrs. All wells were then washed once with 1xPBS, and stained with 0.5% crystal violet solution for 20min while placed on a rocker. Plates were then washed three times with distilled water and dried overnight and resuspended with methanol and quantified with a plate reader at 540nm wavelength.

### Clinical Trial

This study was approved by the Columbia Institutional Review Board (IRB-Q3709)

### Animal studies

All mouse experiments were conducted using protocols approved by the Institutional Animal Care and Use Committee (IUCAC) at the Icahn School of Medicine at Mount Sinai.

- ***Transgenic and xenograft mouse models*** FVB/N-Tg(MMTVneu)202Mul/J transgenic mice were purchased from Jackson laboratory and bred. Six to eight week old NOD.Cg-*Prkdcscid Il2rgtm1Wjl/*SzJ mice were also obtained from Jackson laboratory. Mice were subcutaneously injected under each flank with 10×10^6 MDA-MB 453 and MDA-MB-436 cells resuspended in equal volume of respective media and matrigel matrix (Corning Ref #354234). Treatment began when tumors reached a volume of 100-150mm3 for both transgenic and xenograft mice. A minimum of 6 mice per cohort was used in the study.
- ***Drug administration*** Ricolinostat, obtained from Acetylon Pharmaceuticals, and was administered for 6 days a week at 50mg/kg. Paclitaxel was obtained from the Mount Sinai Hospital and administered at 6mg/kg twice a week. Both treatments were given intraperitoneally at a volume of 200ul. Tumors were measured twice a week for the duration of the experiment using the ellipsoid volume formula 1/2 × L × W × H. At experimental endpoint, tumors were collected, formalin fixed and flash frozen. Statistical differences were evaluated with one-tailed t test (n >= 6 per cohort).

### Immunohistochemistry

Immunohistochemistry was performed on formalin-fixed paraffin-embedded (FFPE) tumor tissue sections by the Neuropathology Brain Bank Core at Mount Sinai. All Slides were sectioned, mounted, and stained for hematoxylin and eosin, Ki-67(Abcam #ab15580 1:200) and c-caspase 3(Cell signaling #9664s at 1:50). All slides were processed using Ventana Benchmark XT machine by the core facility.

### Statistical analysis of experimental data

Data on experimental graphs related with cell lines and animal studies are represented as mean ± standard deviation unless specified otherwise. Results were analyzed by Student’s t-test and a p-value below 0.05 was considered statistically significant.

### Transcriptomics data collection and processing

- ***Cancer cell lines***. The RNA-seq transcripts per million (TPM) data of 1,165 cell lines representing 29 cancer types from the Cancer Cell Line Encyclopedia (CCLE) project, together with cell line annotations and gene dependency scores were downloaded from the portal of the Dependency Map (DepMap) project (https://depmap.org/portal, version: Public 19Q1).
- ***Transgenic mouse models of breast cancer***. The gene expression profiles of 385 samples from 27 genetically engineered mouse models (GEMMs) of breast cancer and 2 normal mammary tissues were collected in a study by Dr. Charles Perou and downloaded from the Gene Expression Omnibus (GEO) (GSE42640, Agilent Technology gene expression microarray platforms, 22K, 4×44K or 4×180K
- ***The Cancer Genome Atlas (TCGA)***. The TCGA RNA-seq data at both isoform and gene levels for 32 human primary cancer types including breast cancer were extracted from the QIAGEN OncoLand.
- ***The Molecular Taxonomy of Breast Cancer International Consortium (METABRIC)***. We downloaded the normalized METABRIC gene expression profiles (Illumina HT 12 arrays, N=1,981) from Synapse (https://www.synapse.org/#!Synapse:syn1757063).
- ***Inflammatory breast cancer (IBC)***. The pre-processed gene expression profiles of 63 IBC samples and 134 non-IBC patient samples were collected by an IBC study and downloaded from the GEO (GSE23720, Affymetrix Human Genome U133 Plus 2.0). We normalized the data by quantile method and then removed probe sets with no EntrezGene IDs, resulting in 51,997 probe sets representing 20,517 genes. The gene-level expression values were estimated by taking average the expression of all probe sets for the same gene. The data QC was assessed by the “draw.eset.QC” function in the NetBID software. Two outlier samples (T60 and T61) were identified and removed from subsequent analysis.

### RNA-seq analysis of the clinical trial

- ***FFPE sample collection***. Out of 16 evaluable patients in our clinical trial we were able to obtain 11 FFPE biopsy samples before entering the clinical trial.
- ***Data generation***. RNA samples were isolated using the RNeasy Mini Kit (QIAGEN) and subjected to RNA-seq sequencing. The sequencing was performed by Illumina HiSeq 2500 with ∼25 million SE-100 reads at the Genomics Core Facility at Mount Sinai.
- ***Data QC and processing***. Raw fastq data was assessed by FastQC (v-0.11.5) (13). Salmon (v-0.9.1) (14) was used to quantify the expression of transcripts and genes based on the reference genome hg38 (GRCh38) with gene annotation from GENCODE (release 28). Data has been deposit in the GEO portal (GSE148623).
- ***Batch effect removal***. The RNA-seq experiments were performed in three batches based on sample receival time. We then used the “removeBatchEffect” function in the limma R package (v-3.42.2) to remove the batch effects.

### Integration of human breast cancer transcriptomics data from different sources

Gene expression profiles from four datasets of human breast cancer, including TCGA, METABRIC, IBC, and RNA-seq data from our clinical trial, were merged by using the genes shared across all four datasets. Batch effects were detected by NetBID QC and removed by limma as described in the batch removal of clinical trial RNA-seq data.

### Next-generation HDAC6 breast cancer regulon inference

In order to reconstruct HDAC6 regulon with high accuracy and applicability across all breast cancer subtypes, gene expression profiles was extracted from each of the 9 breast cancer subtypes, including 3 (HR+/HER2-, HER2+ (HR+ and HR-), triple negative breast cancer or TNBC) defined by IHC assays of HR and HER2, and 6 (Luminal A, Luminal B, HER2^*overexpressed*^, Claudin^*low*^, Basal-like and Normal-like) defined by PAM50, from over 3,000 primary breast cancer samples from TCGA and METABRIC. We then used SJARACNe, a scalable software tool for gene network reverse-engineering from big data, to reconstruct signaling networks for each of the 9 breast cancer subtypes.

### HDAC6 cancer type–specific regulon inference

TCGA dataset included RNA-seq profiles representing 32 human primary cancer types. In addition to the new HDAC6 regulon for breast cancer as described above, we used the TCGA RNA-seq data as detailed in the previous section and reconstructed HDAC6 regulons for each of the other 31 cancer types by using the SJARACNe algorithm. The parameters were configured the same as used to generate HDAC6 breast cancer regulons.

### HDAC6 score inference by NetBID

Given a gene expression profile in a cohort of study, we calculated the raw HDAC6 score that summarized the activity of the HDAC6 regulon by using the “cal.Activity” function with the method of “mean” in NetBID software. For the HDAC6 score of human breast cancer samples, we used the new HDAC6 regulon for breast cancer. For HDAC6 score across the 31 TCGA cancer types, we used the cancer type matched HDAC6 regulon. To calculate the HDAC6 score in GEMMs of mouse breast cancer, we used the human gene ids transferred based on the human/mouse homology map that was done in the breast cancer GEMM study.

### HDAC6 score inference by VIPER (Virtual Inference of Protein-activity by Enriched Regulon analysis)

HDAC6 relative protein activity based on our next-generation HDAC6 breast cancer regulon was inferred by the VIPER algorithm implemented in the DarwinOncoTarget test, which has been approved by the NYS Department of Health CLIA/CLEP Validation Unit as an offering in the category of “Molecular and Cellular Tumor Markers for Oncology” (Neal, Michael, Assay Validation Review, Wadsworth Center, NY State Department of Health, PFI: 7313, Project ID: 63859, March 8, 2019).

### Overlaps of cancer type–specific HDAC6 regulons

The HDAC6 regulons of 32 TCGA cancer types inferred by SJARACNe, as described above, were summarized in Supplementary Table 8. The regulon overlapping statistics, including the size of overlapped regulon genes and the p-value of Fisher’s Exact test was summarized in Supplementary Table 9. The scatter plot was made by R package ggplot2 (v-3.3.0).

### Progression free survival (PFS) analysis against HDAC6 score in the clinical trial

The PFS in month was calculated from the dates of the trial assignment and disease progression or last follow-up by the “difftime” function in lubridate (v-1.7.8). The PFS analysis of the clinical trial against HDAC6 score, including the COX model fitting and Kaplan–Meier plot, was performed by using the R package survminer (v-0.4.6) (25). The HDAC6 score high and low patients were defined at the cutoff of the mean of HDAC6 scores.

### Receiver operating characteristic (ROC) curve analysis of HDAC6 score in the clinical trial

The ROC curve analysis to evaluate the performance of HDAC6 score in predicting clinical response of patients, including statistics and plot, was performed by using the R package pROC (v-1.6.2) (26). We used - 0.36 as the cutoff of HDAC6 score from the ROC analysis to define HDAC6 high and low patients.

## SUPPLEMENTARY FIGURE LEGENDS

**Supplementary Figure S1.**
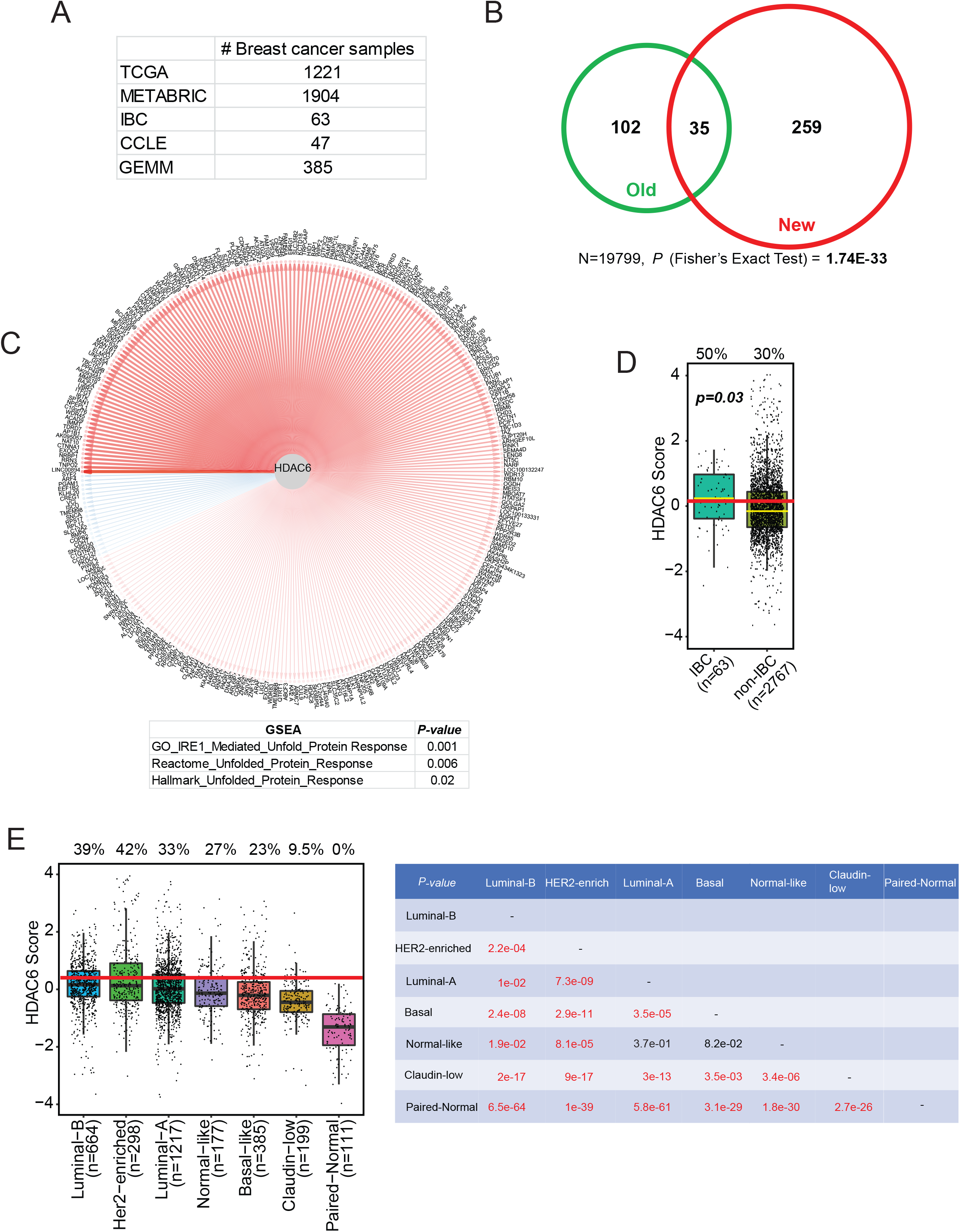
HDAC6 score in BC. (A) The number of samples and data set of origin used to evaluate the HDAC6 regulon. (B) Overlap of old and new HDAC6 regulons. (C) The network plot of HDAC6 and updated HDAC6 regulon. Edge width is corresponding to the correlation strength measured by mutual information. Red and blue edges indicate positive and negative correlations between HDAC6 and each of the regulon genes. The table shows the GSEA of the genes in the regulon showing its association with unfolded protein response. (D) New HDAC6 score comparing IBCs vs non-IBCs. (E) HDAC6 scores of all BCs from TCGA and METABRIC divided into molecular subtypes. The red line represents the median of the HDAC6 scores in IBC samples and the numbers over each whisker plot indicate the percentage of samples over this value in each clinical subtype. The p-values comparing the HDAC6 scores of all the groups are also shown.

**Supplementary Figure S2.**
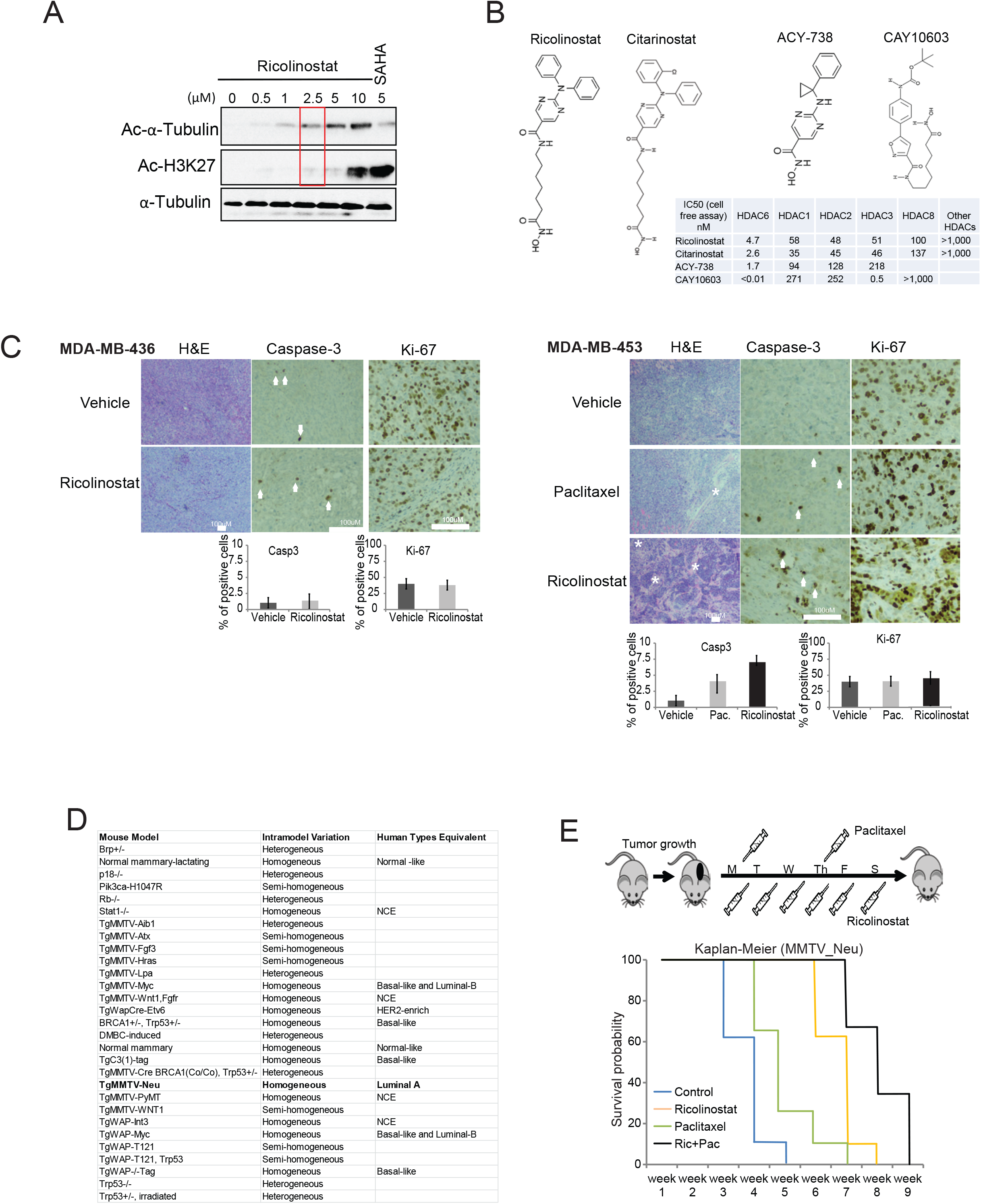
Anticancer activity of HDAC6 inhibitors. (A) The western blot shows the titration of ricolinostat to identify an effective dose (accumulation of Ac-α-Tubulin) without off-target effects in class-I HDACs (accumulation of Ac-H3K27). SAHA is used as a control Pan-HDAC inhibitor. (B) Chemical structure of the different HDAC6 inhibitors used in Figure 1E. (C) Histological intratumoral evaluation of H&E, Caspase-3, and Ki-67 in tumor samples from Figure 1B. Quantification is also shown in bar graphs. Notice that the combo treatment (Pac+Ric) is not shown because all tumors regressed with this treatment. (D) The list shows all the transgenic mouse models evaluated by the HDAC6 score in Figure 2 C and D and indicates their correlation with human BCs (E) Kaplan – Meier graphic showing the survival of the MMTV tumors in Figure 2E.

**Supplementary Figure S3.**
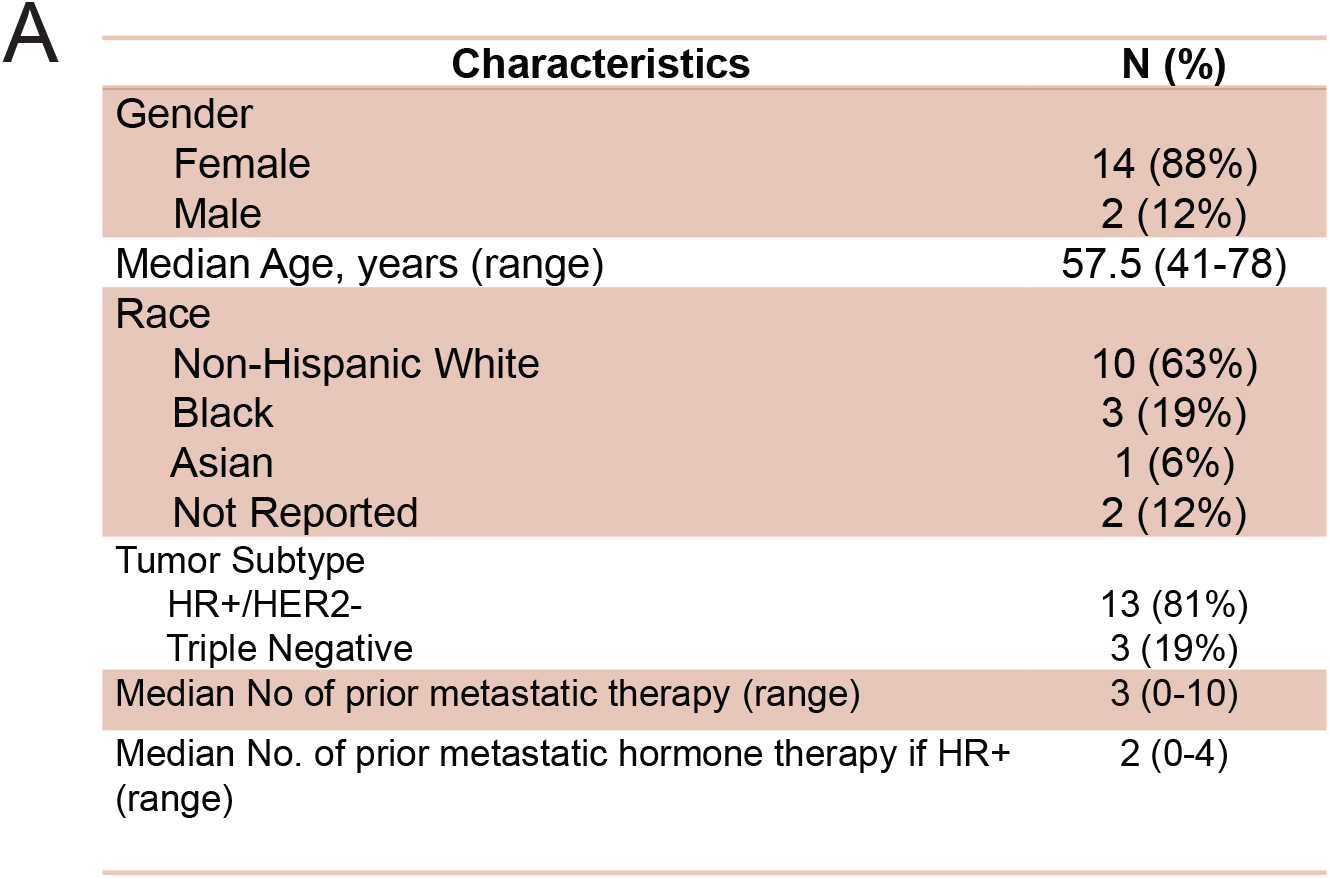
Characteristics of the evaluable patients enrolled in the clinical trial.

**Supplementary Figure S4.**
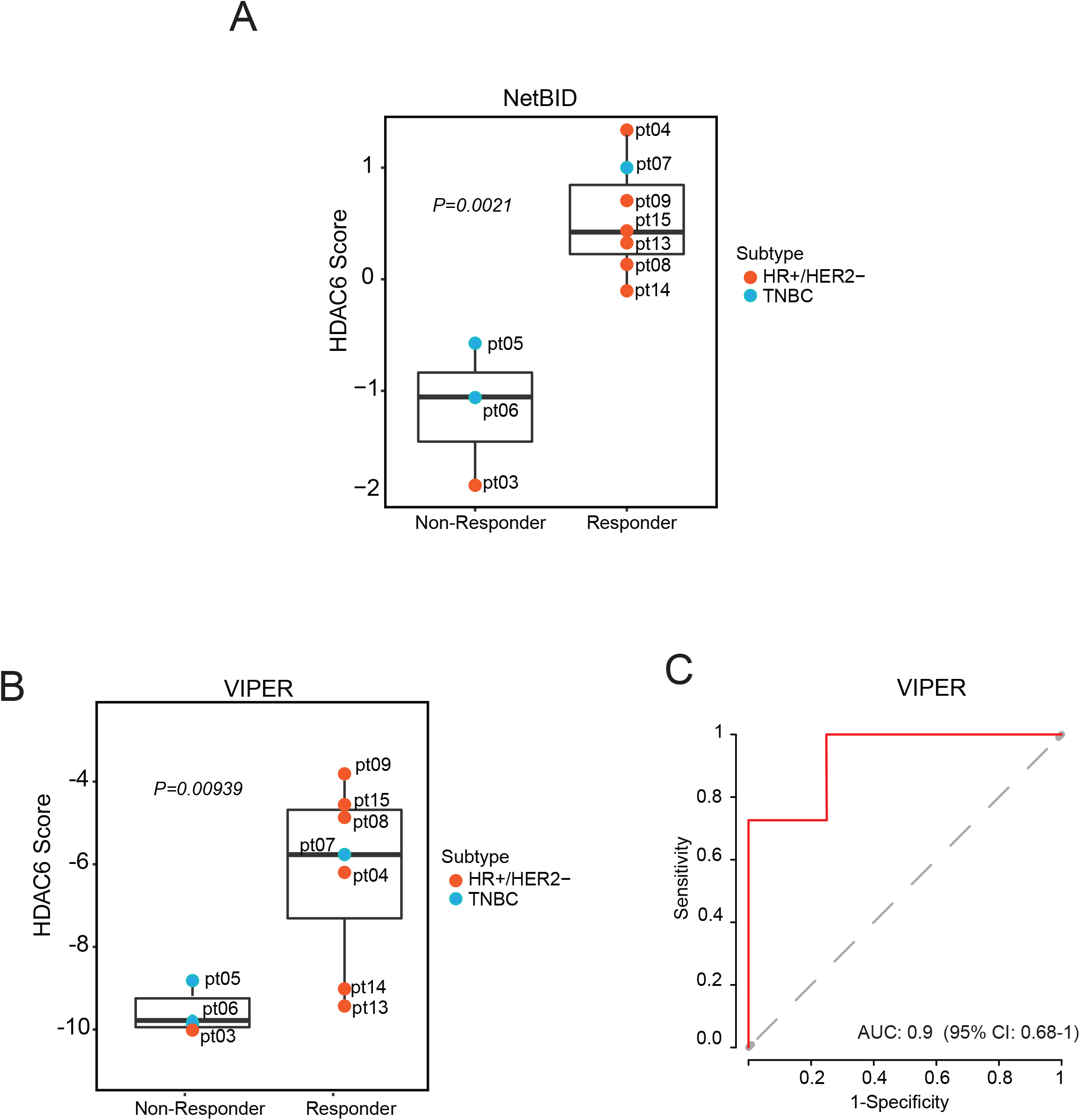
Biomarker evaluation of HDAC6 score calculated by using the NetBID and VIPER algorithms in the phase Ib trial. The figure shows the similarities between the HDAC6 scores inferred by NetBID (A) and VIPER (B) in responders and non-responders, and (C) ROC curve plot of HDAC6 score inferred by VIPER (similar plot for NetBID is in Figure 3D).

**Supplementary Figure S5.**
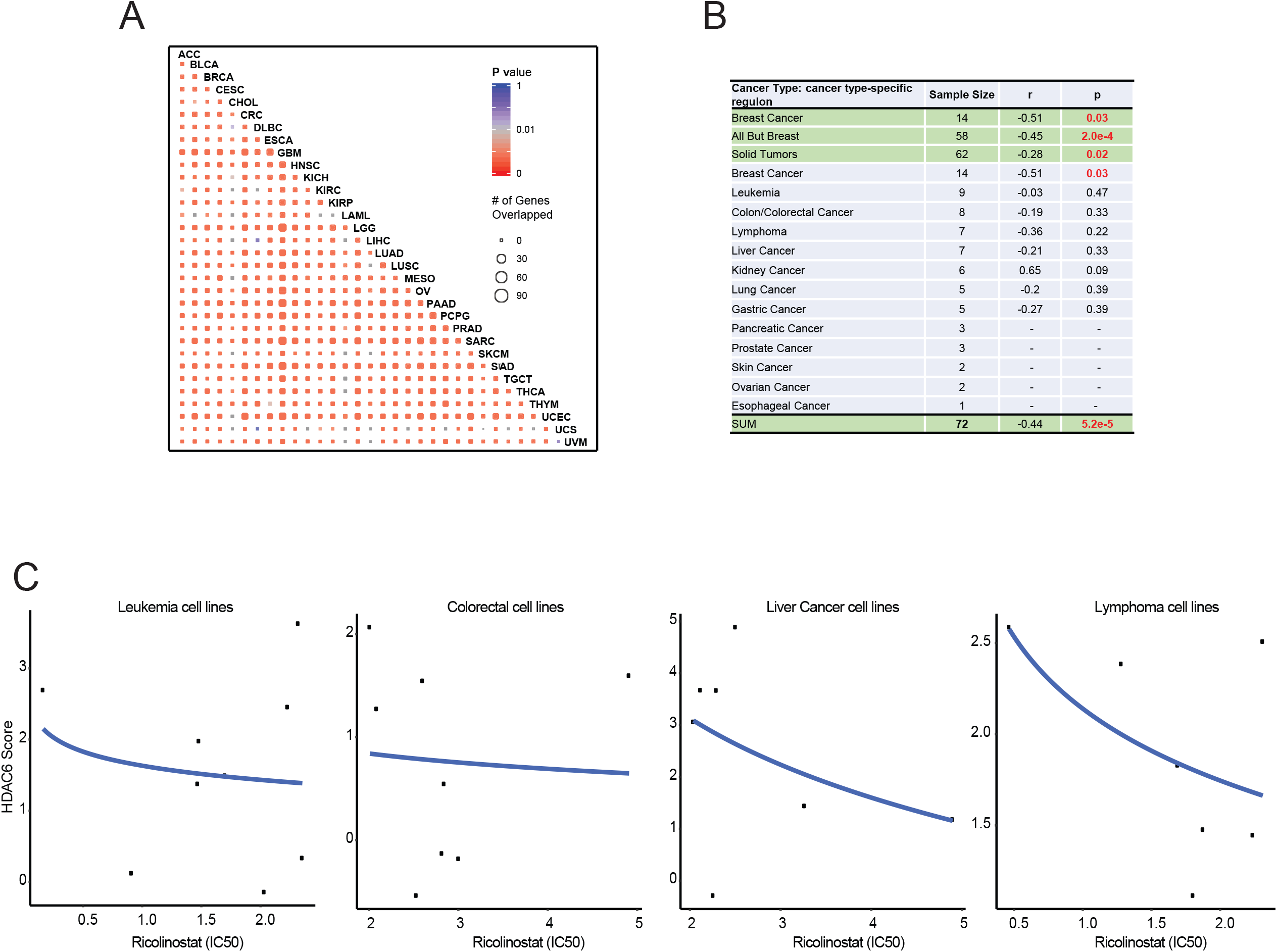
HDAC6 scores in other human cancers. (A) Correlation between HDAC6 regulon in different tumor types. LAML: Acute Myeloid Leukemia; LIHC: Liver Hepatocellular Carcinoma; UVM: Uveal Melanoma; KIRP: Kidney Renal Papillary Cell Carcinoma; PCPG: Pheochromocytoma and Paraganglioma; DLBC: Diffuse Large B-Cell Lymphoma; ACC: Adrenocortical Carcinoma; UCS: Uterine Carcinosarcoma; KIRC: Kidney Renal Clear Cell Carcinoma; THYM: Thymoma; CHOL: Cholangiocarcinoma; SKCM: Skin Cutaneous Melanoma; CRC: Colorectal Carcinoma; LGG: Brain Lower Grade Glioma; UCEC: Uterine Corpus Endometrial Carcinoma; KICH: Kidney Chromophobe; MESO: Mesothelioma; TGCT: Testicular Germ Cell Tumors; GBM: Glioblastoma; PRAD: Prostate Adenocarcinoma; CESC: Cervical Squamous Cell Carcinoma and Endocervical Adenocarcinoma; BLCA: Bladder Urothelial Carcinoma; SARC: Sarcoma; OV: Ovarian Serous Cystadenocarcinoma; THCA: Thyroid Carcinoma; ESCA: Esophageal Carcinoma; BRCA: BC; LUAD: Lung Adenocarcinoma; HNSC: Head and Neck Squamous Cell Carcinoma; PAAD: Pancreatic Adenocarcinoma; STAD: Stomach Adenocarcinoma; LUSC: Lung Squamous Cell Carcinoma. (B) List with all the cell lines evaluated by dose-response to ricolinostat and HDAC6 scores. The correlation (r) and p-value (p) between the response to ricolinostat and HDAC6 scores are also shown. (C) Graphic showing the correlation between the HDAC6 score and the response to ricolinostat in individual cell types (only tumor types with cell lines n>6 are shown).

## Notes

**DISCLOSURES** - MY and SJ were Acetylon employees when this project was initiated. - This research has been partially supported by a sponsor research agreement with Acetylon. - JY was a consultant of the computational analysis for the Phase Ib trial 2015-2017. - MJA is Chief Scientific Officer and equity holder at DarwinHealth, Inc., a company that has licensed some of the algorithms used in this manuscript from Columbia University. AC is founder, equity holder, consultant, and director of DarwinHealth Inc., a company that has licensed some of the algorithms used in this manuscript from Columbia University. Columbia University is also an equity holder in DarwinHealth Inc.

### Competing Interest Statement

DISCLOSURES
- MY and SJ were Acetylon employees when this project was initiated.
- This research has been partially supported by a sponsor research agreement with Acetylon.
- JY was a consultant of the computational analysis for the Phase Ib trial 2015-2017.
- MJA is Chief Scientific Officer and equity holder at DarwinHealth, Inc., a company that has licensed some of the algorithms used in this manuscript from Columbia University. AC is founder, equity holder, consultant, and director of DarwinHealth Inc., a company that has licensed some of the algorithms used in this manuscript from Columbia University. Columbia University is also an equity holder in DarwinHealth Inc.

### Clinical Trial

NCT02632071

